# A Community-Engaged Public Health Research and Outreach Program for Migrant and Racialized Workers in Meat Processing to Mitigate COVID-19 Inequities

**DOI:** 10.1101/2025.05.28.25327793

**Authors:** Gabriel E. Fabreau, Eric Norrie, Linda Holdbrook, Minnella Antonio, Mohammad Yasir Essar, Michael Youssef, Adanech Sahilie, Mussie Yemane, Edna Ramirez-Cerino, Nour Hassan, Rabina Grewal, Zahra Hussain, Deyana Altahsh, Olivia Magwood, Ammar Saad, Maria Santana, Aleem Bharwani, Ingrid Nielssen, Samuel T. Edwards, Denise Spitzer, Annalee Coakley, Kevin Pottie

## Abstract

**Objective:** COVID-19 has disproportionately impacted migrant workers in meat processing industries causing mass outbreaks and fatalities. Implementing community based participatory research (CBPR) methods may increase public health engagement, but developing the prerequisite trust required is hindered during a public health crisis.

**Methods:** We used CBPR methods to recruit, train and integrate six community scholars representing various racialized ethnocultural minorities into public health research and outreach operations. We present an organizational case study of their experiences across multiple Canadian meat plants affected by mass COVID-19 outbreaks. We used administrative documents to describe the project setting, training, and roles across research and vaccine operations between March 2020 and December 2022. Scholars then completed reflexivity activities using narrative analysis to summarize their experiences and impacts on themselves, migrant workers, and their communities. Finally, we integrated our data through scholars’ reflections to investigate how their narrative analysis was reflected in the administrative, quantitative and time series data.

**Findings:** We summarize three study phases; 1) Scholars’ recruitment and training; 2) early community engagement; 3) community outreach vaccinations. After Scholars’ team integration, initial worker study recruitment attempts failed due to mistrust and fear of employer reprisals. Scholars built trust among workers playing key roles in nine onsite meat plant occupational and community outreach COVID-19 vaccine clinics. successfully surveyed 191, and interviewed 43 workers in seven primary languages across eleven meat plants between January 2021 and February 2022. Scholars described their roles, successful outreach strategies, learnings, prerequisite skills, and intimate interactions that contributed motivation and meaning. Key insights included empathetically validating workers’ experiences, translating stories into advocacy, and the importance of community presence combining public health research and outreach.

**Conclusion:** During public health crises, community-academic-healthcare partnerships can rapidly implement multicultural CBPR strategies to effectively engage migrant workers concurrently in both research and public health outreach.

**Funding:** Canadian Institutes of Health Research (CIHR Application no. 469206)

**Key Messages:** *What is already known on this topic?:* The COVID-19 pandemic disproportionately impacted migrant workers in meat processing industries, leading to significant outbreaks, poor health outcomes, and fatalities across multiple high-income countries. Existing literature highlights the challenges of engaging these workers in public health research and outreach operations due to structural barriers, precarious economic and immigration statuses, and mistrust towards health authorities. Community-based participatory research (CBPR) methods can overcome these barriers; however, they fundamentally depend on developing trust between academic and healthcare partners and migrant workers, which is very difficult during public health crises such as mass COVID-19 outbreaks in meat processing facilities.

*What this study adds?:* This study introduces and evaluates a novel, rapidly developed community-based participatory research (CBPR) program that integrated six community leaders called ‘Community Scholars,’ representing various racialized ethnocultural minorities, into public health research and outreach operations concurrently. It details how community scholars were trained and integrated into teams to engage migrant workers in research and vaccine outreach operations. The study outlines the program’s failures, successes, reflections, and key learnings, to overcome traditional participation barriers.

*How this study might affect research, practice or policy?:* These findings suggest that employing CBPR methods rapidly with active community involvement can synergistically enhance engagement and trust among migrant workers across both public health research and operations. This study provides insights that may serve as a blueprint for similar contexts, informing future public health strategies and policies to better manage crises involving socially vulnerable migrant populations. It emphasizes the potential of community-driven approaches to bridge gaps in public health research, practice and policy, particularly during emergencies.

## 1. Introduction

The COVID-19 pandemic disproportionately impacted frontline workers in meat processing facilities, leading to numerous outbreaks, community spread, and fatalities.(1–4) The nature of these workplaces – large, closely packed workforces operating in poorly ventilated, physically demanding, noisy and humid conditions – create environments conducive to rapid viral transmission.(1,5–9) In high income countries, these industries predominantly employ migrant workers, a diverse group encompassing immigrants, refugees, temporary and undocumented foreign workers(10) and racialized national staff.(7,11,12) These workers face numerous structural barriers that increase infection risks and limit their access to health services(7,13,14) including language and cultural differences, communal living, low-quality housing, transportation barriers that require carpooling and public transport, and financial insecurity.(3,4,6,8,14) The COVID-19 pandemic exacerbated these challenges, with repeated outbreaks, lock-downs, and emergency public health measures.(1,15) These factors, combined with institutional mistrust and fear of employer reprisals have hindered efforts to engage migrant workers in health-related research and public health campaigns, making migrant workers’ lived experiences and perspectives largely unheard.(12,13,16)

Community-based participatory research (CBPR) methods offer potential solutions to overcome these barriers and provide tools for meaningful representation, inclusion, and engagement of marginalized communities such as migrant workers in public health research and outreach campaigns.(17) These methods fosters reciprocal trust, codesign, and minimized power hierarchies between researchers and community members; however, community participation exists on a spectrum of control, with researchers on one end and community members on the other.(18–21) The knowledge co-produced by community-research partnerships aims to reduce health inequities and improve health system and public health operations.(18,22–25) The World Health Organization (WHO) recently highlighted migration health research as a global priority, specifically to improve knowledge on response to health emergencies, and inclusive research partnerships as critical to advancing the Global Action Plan on promoting the health of refugees and migrants, 2019–2030, and to achieving the health-related Sustainable Development Goals.(25,26) However, these methods are often pursued sequentially not concurrently (i.e., research first), and fundamentally require time to build trust, identify, train, and integrate migrant community members into existing research and operational teams – time that public health emergencies do not afford.(19) Mass COVID-19 outbreaks in meat processing facilities necessitated urgent mobilization of both participatory public health research and outreach operations without time to develop meaningful community relationships, nor to conduct research prior to implementing emergency public health operations.

We undertook an investigation of meat processing plants in western Canada, two of which experienced among the largest COVID-19 outbreaks in North America (April-July 2020).(1,11,27,28) As part of this broader study, we employed CBPR principles to engage with migrant workers and to understand if a meaningful CBPR program could be rapidly co-developed under pandemic conditions integrated across both public health research and urgent outreach operations. To investigate this, we conducted an organizational case study of CBPR methods concurrently developed for public health research and outreach during the first two years of the COVID-19 pandemic.(20,29) We present this study aiming to provide guidance for other jurisdictions facing similar challenges engage with migrant workers and respond to future public health emergencies.

## 2. Methods

### 2.1 Community Based Approach and Study Context

This study pursued a community based participatory research (CBPR) approach to investigate the pandemic’s impact on migrant workers across multiple meat processing plants in western Canada between March 2020 and March 2022. We present our experiences – the false starts, failures, successes, reflections, and key lessons from our “Community Scholars” (CSs) program, which integrated community leaders representing various minoritized ethnocultural groups into our research team and public health outreach operations. We detail the processes required to recruit, train, integrate, and empower a Community Scholar team to concurrently conduct research and facilitate COVID-19 vaccine outreach among migrant workers during a public health emergency.(20,29) Finally, we present the CSs’ personal reflections, impacts, lessons learned, and recommendations.

We adopted Sanchez et al.’s CBPR implementation framework that emphasizes community-based, participatory approaches to improve health conditions by reducing social and structural determinants and injustices.(17,21,30) We define meaningful engagement as per O’Reilly de Brun et al.’s definition as, “an experience of research [and operational] engagement that is collegial, inclusive and active for participants and which enables their perspectives to emerge clearly in research outcomes”.(31) We used the International Organization for Migration’s definition of *migrants* as, “a person who moves away from his or her place of usual residence, whether within a country or across an international border, temporarily or permanently, and for a variety of reasons”.(10)

We implemented our community-academic-health system partnerships early in the pandemic during public health restrictions through Canada’s initial mass COVID-19 vaccination campaign (Figure 1a). Throughout the study, our team was concurrently involved in public health research and operational efforts that included, disseminating health information, facilitating healthcare and social services access, and implementing mass occupational COVID-19 vaccine outreach clinics at various meat processing facilities and in socially deprived urban neighborhoods with high proportions of migrants and racialized peoples.(32,33) These operational roles depended on partnerships between the provincial health system, and various stakeholders and entities detailed previously.(32,34)

**Figure 1a:**
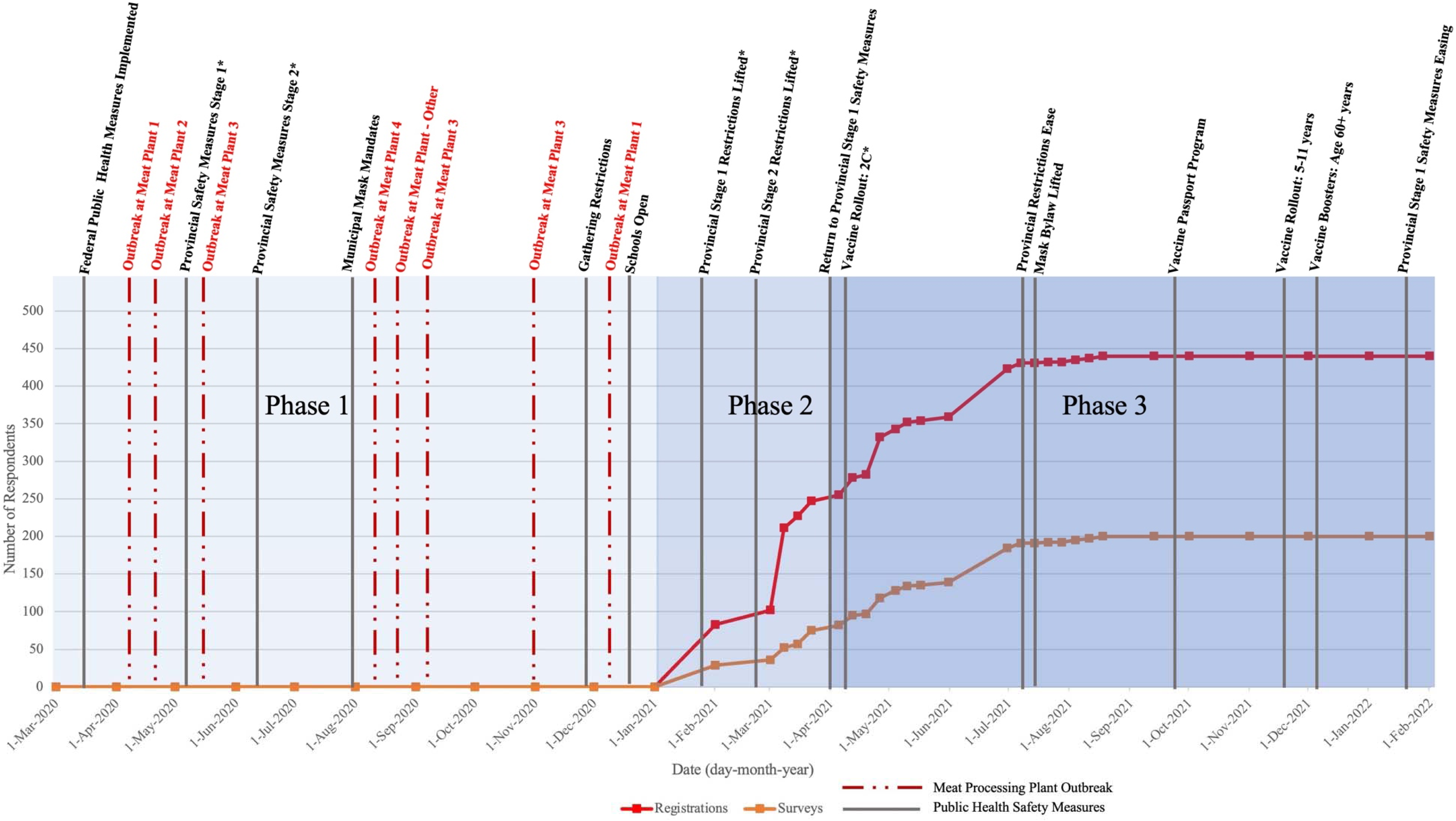
Number of registrations and survey respondents across three phases of study recruitment, within the context of public health measures & meat plant outbreak data from March 2020 to February 2022. *Stages 1 and 2 refer to the stages of the provincial re-opening plan to loosen public health measures: Stage 1: Some businesses and services re-opened, continued physical distancing, public face masks encouraged. Stage 2: Additional businesses and services re-opened; continued physical distancing, public face masks encouraged, increased gathering limits. ** Vaccine 2C: Within provincial Covid-19 immunization schedule, the 2C category prioritized meat plant workers (and other high-risk employees) to receive vaccinations. eTable 1 presents additional information regarding Alberta’s COVID-19 public health measures and immunization schedule.

### 2.2 Community Scholars Selection and Training

Community Scholars (CSs) and study team members were initially introduced during the public health response to mass COVID-19 outbreaks in various meat plants (Figure 1a).(32) Each CS volunteered or worked with different community organizations to support migrant meat plant workers during these mass outbreaks. We recruited CSs from six different minoritized ethnocultural communities. Of these, five completed the program and study, representing local migrant worker populations. The CSs received research training previously used in other contexts aimed at building community health research competency and technical skills in public health research (35,36)

### 2.3 Study Tools, Co-Design, Data Sources, and Study Timeline

The CS team was integrated into the research team upon study launch and was involved in all study aspects, including co-design, refinement, study recruitment, data collection, analysis, interpretation, and knowledge mobilization. The study team and CSs co-designed operational research team documents, tools, CS training materials, and reflexivity exercises. Our data sources include: 1) Community Scholar (CS) training curriculum and materials; 2) research instruments and tools (surveys and interviews); 3) study recruitment strategies, materials and outreach tracker; and, 4) CS reflexivity exercise. We conducted this research across three study phases; 1) CS recruitment, training and integration (March 2020–December 2020; 2) unsuccessful meat processing worker recruitment (January–March 2021); and, 3) outreach vaccinations, successful meat processing worker recruitment, and CS reflections (April 2021– March 2022).

### 2.4 Community Scholar Reflexivity

We conducted reflexivity activities, inviting CSs to reflect on their experiences, their impacts on community members, the CS program’s key learnings, and their recommendations for future public health research and outreach operations. Each CS individually documented their reflections, collaboratively highlighted reflections they felt were most impactful and representative of their lived experiences, then collectively summarized their key learnings.

### 2.5 Data Analysis and Integration

We integrated our data sources using a sequential mixed-methods design that began with quantitative and document analysis, followed by qualitative narrative analysis of CS’ reflections to investigate the relationship between the CS program, migrant worker study recruitment, and vaccine outreach efforts.(37) Two study team members utilized narrative analysis to integrate the CS reflections into the following codes: 1) Narrative summary; 2) Orientation - who, when, where?; 2) Complicating Action - what happened?; 3) Evaluation - CS interpretation; 4) Result - key themes and learnings.(32,38) The entire study team then pursued a collective sense-making approach to review the initial codes, finalize the narrative analysis, and summarize the key learnings.(20,39) The study team integrated these qualitative findings with CS program administrative document analysis, and quantitative data that summarized study recruitment, including registrations, completed surveys and interviews among meat plant workers. Quantitative time series data regarding study participant recruitment, registrations, completed surveys and interviews were collected, managed, and analyzed using REDCap electronic data capture tools(40) and Microsoft Excel (Redmond, Washington, USA). We analyzed qualitative data with NVivo Version 12 data analysis software (QSR International, released March 2018; Burlington, Massachusetts).(41) The study was approved by the University of Calgary research ethics board (REB20-1153). To summarize the study, we utilized the Good Reporting of a Mixed Methods Study (GRAMMS) and Guidance for Reporting Involvement of Patients and the Public (GRIPP2) reporting guidelines.(42,43)

## 3. Results

### 3.1 Narrative Summary

Community Scholars were initially trained to engage meat plant workers in research; however, their role expanded to public health advocacy, operations, and COVID-19 vaccine outreach. Early study recruitment failed repeatedly due to the heightened workplace precarity, and fear amplified by COVID-19 outbreaks. CSs persevered, leading community engagement even during difficult public health restrictions. By spring 2021, the study team proposed and co-led onsite occupational COVID-19 vaccination outreach activities at several surrounding meat plants (Figure 1b). By leveraging community relationships and the study teams’ onsite presence, these outreach clinics saw both high worker vaccination rates and increased research engagement.(32)

**Figure 1b:**
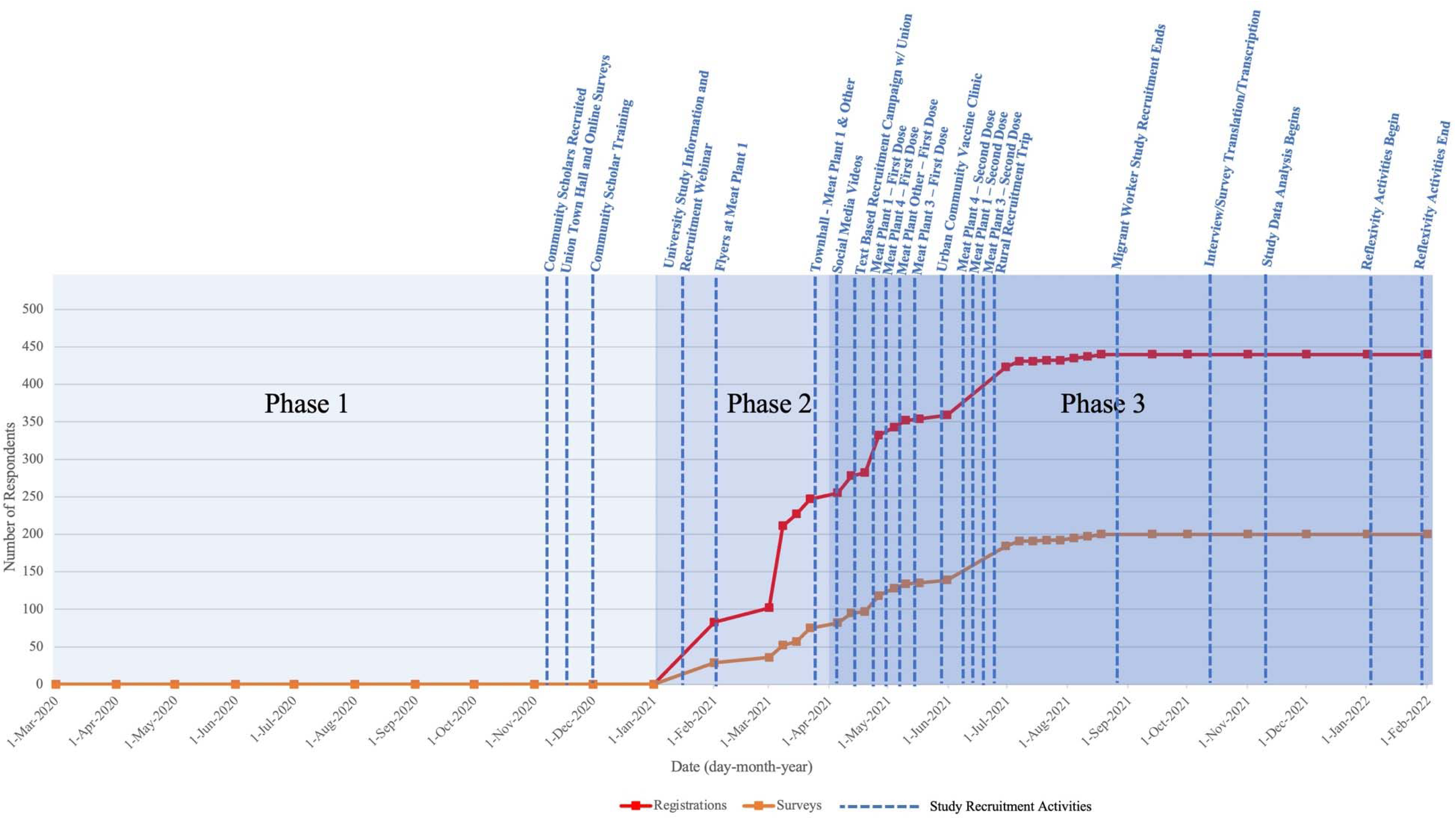
Phase 1 Community Scholars public health research recruitment and outreach activities from March 2020 to February 2022.

### 3.2 Orientation: Meat Processing Plants, Community Scholars, and Communities

The CS team recruited workers from 11 meat processing plants across western Canada whose workforce comprised largely of racialized migrant workers, along with other national staff. These plants were classified as essential food services and remained mostly operational despite large COVID-19 outbreaks.(11)

#### Phase 1: Community Scholar recruitment and engagement

Six CSs led community-engagement efforts. They spoke eight primary languages: English, Amharic, Oromo, Spanish, Tagalog, Arabic, Tigrinya, and Dinka. Table 1. summarizes Community Scholars’ demographics, training, education, and community engagement experiences. Early after the pandemic was declared in Canada and the first mass meat processing plant outbreaks (Figure 1a), CSs either volunteered or worked for newcomer and migrant serving agencies that were integrated into the provincial health system’s emergency response. Scholars conducted contact tracing, provided social support such as food and emergency housing to help infected workers isolate, helped translate and navigate complex social programs, and delivered public health information. Thus, CSs became familiar with meat processing plant workers and their communities.

**Table 1:** Community Scholars Characteristics.

| Pseudonym | Age Band (5-yr) | Languages spoken | Education/ Occupational Experience | pre COVID-19 Community Engagement & Leadership Experiences |
| --- | --- | --- | --- | --- |
| Participant A | 50-55 | Tigrinya, Amharic, English | Economist; MSc, Agricultural Economics and Agribusiness, MA and BA Honors in Economics | Former President of Community Association |
| Participant B | 35-40 | Amharic, Tigrinya, English | BA in Education and Teaching diploma, Certified in DBt, Trauma Informed Care and Community Mental Health | Founder of immigrant serving agency, City Police Anti-Racism Action Committee |
| Participant C | 30-35 | Spanish, English | MD; International Medical Graduate | Member of Health & Wellness team - [Province] organization for internationally Trained Physicians |
| Participant D | 35-40 | Arabic, English | MD; International Medical Graduate | Member of Health & Wellness team - [Province] organization for internationally Trained Physicians |
| Participant E | 25-30 | Tagalog, English | BComm, certificate in Social Innovation | Volunteer with various ethnocultural community organizations. Community broker during COVID-19 pandemic. Community engagement coordinator for local businesses |

#### Phase 1: Community Scholar Personal Impact and Motivations

Community Scholars shared their motivations to participate in research, deeply influenced by their experiences supporting migrant workers:

> *“When COVID hit, the East African community was severely impacted economically, socially, and emotionally. I was frustrated and cried so many nights when I got multiple calls from the community members.” – Participant B*

> *“I heard many, different genders, different countries of origin, different meat processing plants, but the story kept repeating itself. The mistreatment, the harassment, the use of power to manipulate…” – Participant E*

These interactions with community members motivated CSs to act and join the research team.

> *“The more people I spoke to the more I felt we needed to do something to help them.” – Participant C*

> *“My frustration led me to mobilize my resources and create a [community] group.” – Participant B*

> *“When the first COVID-19 outbreak happened at [Meat Plant] I was providing support to those affected… When this [research] project started, I knew I wanted to be involved…” – Participant C*

#### Phase 1: Community Scholar training and integration

The CS team training was deployed between November 2020 throughout December 2021 and involved: 1) an introduction to the study’s objectives and population; 2) global health competencies such as cross-cultural communication, advocacy, and leadership (through an e-learning model)(35,36); and, 3) technical research training including research ethics and certification, data security and privacy, informed consent and participant recruitment, survey data collection, qualitative interview facilitation, and analysis methods. In January 2022 the CS team began a formal 1-year university-based research certificate training program.(44,45) Community Scholar training and its components are detailed in Figure 1b.

### 3.3 Complicating Action

#### Phase 2: Early study recruitment and community engagement

Despite employing various outreach techniques to encourage research participation among meat plant workers, these early methods were ineffective (Figure 1b), and summarized in Table 2.

**Table 2:**
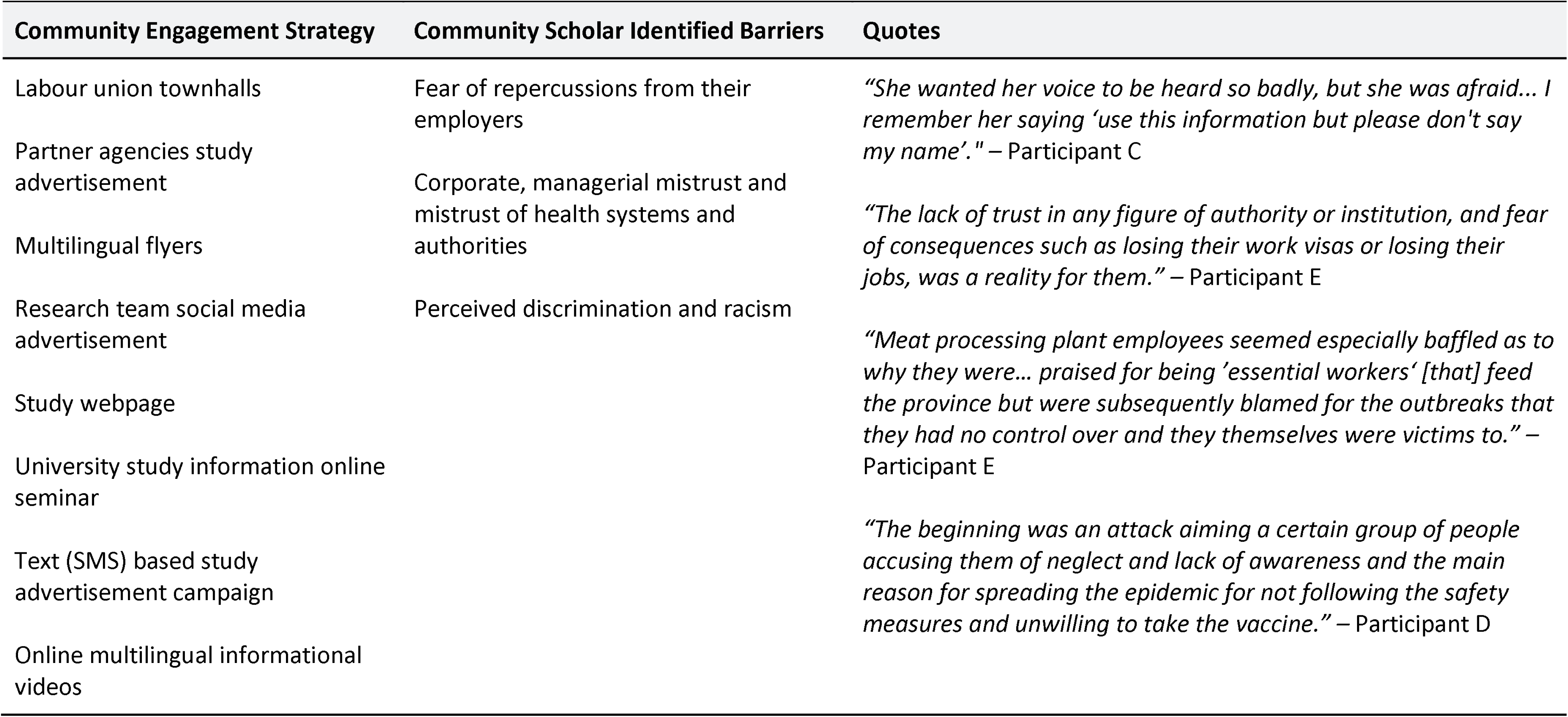
Early and Unsuccessful Meat Plant Worker Engagements.

### 3.4 Community Scholar Evaluation - Reflections

> *“It was like a spy mission to uncover the disaster … this was [our] main role as a community scholar (spy-eye) … we put a spotlight on the government’s weaknesses on handling crisis.” – Participant D*

Scholars reflected on the workers‘ fears, difficult social and working conditions, perceived exploitation, and racial discrimination, but also how for temporary workers, job security and immigration status were tied to employment.

> *“Many felt unsafe and scared going to work every day, some with no choice other than to carpool as they did not have their own vehicles and with many also living in the provided shared housing of their employer. Many meat plant employees had to continue working because their immigration status relied on their ability to work despite how terrifying it was…” – Participant E*

> *“[One worker told me] ‘Last year and prior to that, in 8 hours we used to kill 2,000 to 2,050, but now we are killing 2,200 to 2,300 cows. The job is getting harder…[Meat Plant] has a structure the same like the socialist dictator government we had in Ethiopia’…” – Participant A*

> *“The employees I spoke to say the main reason for the spread of COVID in 2020 and beyond was the working conditions in the meat processing plant. They work on lines, stand shoulder-to-shoulder, making the same cuts over and over.” – Participant A*

> *“Several the employees believe that there is a hierarchy based on race and there is a divide and rule mechanism in the meat processing plants.” – Participant A*

One CS described reflecting on their own immigrant status while supporting a migrant worker who was crying.

> *“I distinctly remember one woman who called me crying, we spoke for two hours, she told me how her job was unbearable and how she had been psychologically harassed by her employers… Her case was full of mistreatment and injustice. It was the first time I heard someone saying, ‘it is like we are slaves in the 21st century’. This was shocking to me being an immigrant myself.” – Participant C*

Scholars reflected on the personal impacts they experienced engaging with migrant workers and their communities.

> *“Being part of the research team inspired and motivated me to continue to support essential workers.” – Participant B*

### 3.5 Resolution - Phase 3: Successful Worker Engagement and Integration with Occupational and Community COVID-19 Outreach Vaccine Clinics

> *“We were experiencing failure, for…interviews and surveys. We didn’t surrender, we continued and ultimately achieved success. ” – Participant D*

Despite early challenges CSs remained highly motivated, completed their training, and pursued multiple engagement strategies. They identified various recruitment barriers and led iterative improvements in recruitment methods to address them (Table 3). Community Scholars became further empowered to lead engagement efforts, using weekly team meetings to discuss engagement activities and track study recruitment (Figure 1b).

**Table 3:**
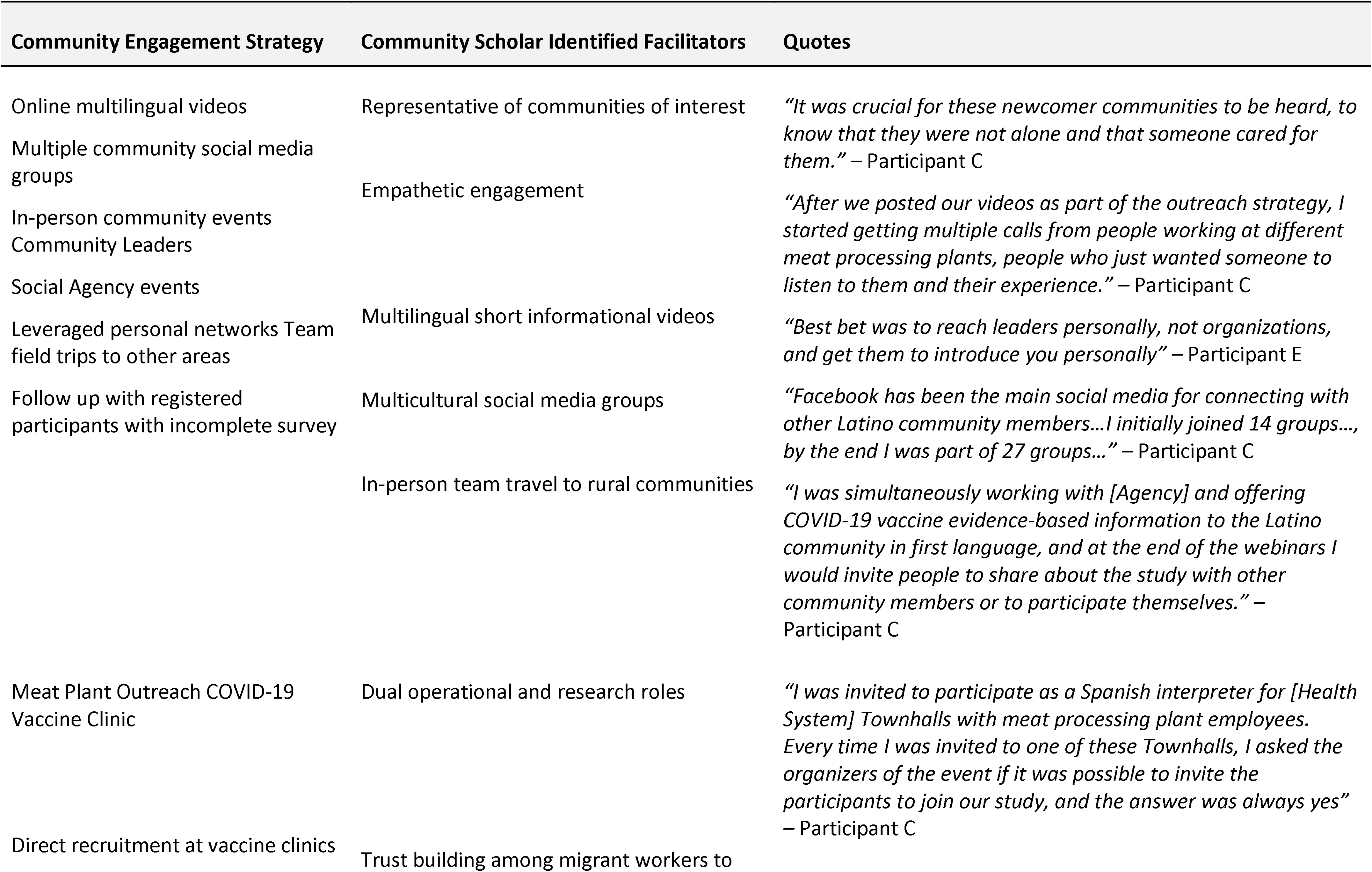
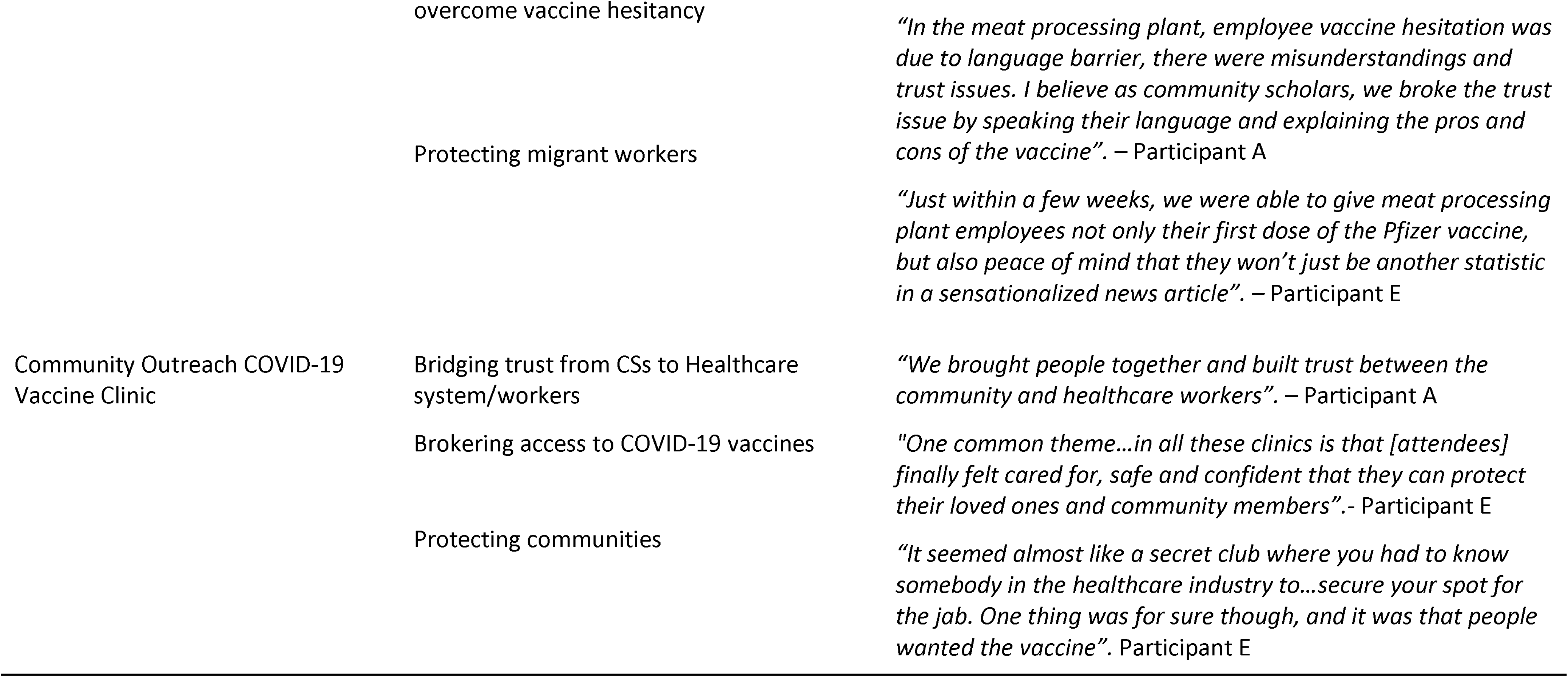
Iteratively improved and successful migrant worker community engagement outreach methods.

Later CSs utilized their operational presence during eight on-site occupational meat plant and one urban COVID-19 vaccine outreach clinics, to facilitate study recruitment. Scholars successfully registered 450 meat processing plant workers, facilitated 191 survey completions (42.4% completion) and performed 43 qualitative interviews across six different primary languages (Figure 1b). Table 3 presents the CS team iteratively improved migrant worker engagement methods and study recruitment integration with COVID-19 vaccine outreach clinics.

Scholars also described their experience brokering access to COVID-19 vaccines for migrant workers and their communities, finding this role particularly important when working with migrant communities who experienced elevated levels of mistrust towards the health system and towards COVID-19 vaccines.

> *“To make matters worse, most Immigrants from East Africa had vaccine hesitancy… Their suspicion comes from their disenfranchisement and isolation from the mainstream system.” – Participant B*

> *“The mobile meat plant clinics were incredibly convenient for the employees as they were able to get their vaccine before or after their shift and cooperation with the employers made the process smooth and easy.” – Participant E*

### 3.6 Key Learnings

> *“We cannot underestimate the power of community and feeling cared for and supported in someone’s life… It is true that we are stronger together, and trust can be built when we show with actions that we care for one another.” – Participant C*

Several key insights emerged from the Community Scholars’ experiences:

1. Validating workers’ experiences and empathetic listening synergistically facilitated both public health research and outreach efforts: Community Scholars underscored the importance of bearing witness to migrant workers’ challenges, grievances, and concerns.

> *“CBPR with essential workers gave them a chance to express and share their lived experiences, creating social connection and meaningful relationships among themselves. As such, the research was more than research, the research involvement was a validating experience for essential workers.” – Participant B*

> *“Before [the] Community Scholars, people were very hesitant to take survey and do interviews. But as soon as Community Scholars joined, people joined the survey and interview because people look and talk like them.” – Participant A*

> *“It was crucial for these newcomer communities to be heard, to know that they were not alone and that someone cared for them.” – Participant C*

2. Translation of stories into advocacy: CSs advocated for workers’ rights, helping them protect themselves and their communities.

> *“We started to give hope to the employees…to learn about their rights and stopped normalizing the way they were treated…we talked to them, they realized that their experience is not every immigrant experience…which encouraged them to speak up.” – Participant C*

3. Community presence in the face of uncertainties: CSs stressed the significance of maintaining a community presence beyond research and to reporting all research findings through continued community dialogue and presence.

> *“I feel much closer to my community, I made many deep and meaningful connections… just keep showing up. Don’t just abandon them. [Keep going] to the BBQs, go to the potlucks. Don’t just take.” – Participant E*

> *“Being at the receiving end of the trust of our community members is both a great honour and a great responsibility. This has been my motivation when I have felt tired or overwhelmed.” – Participant C*

## 4. Discussion

In this study we describe how a community scholar program cultivated trust among migrant workers in meat plants affected by COVID-19 outbreaks, facilitated through empathetic listening, onsite presence, and translation of worker narratives into advocacy during the pandemic. Our findings demonstrate the feasibility of rapidly deploying and refining community-based participatory strategies to engage migrant workers to concurrently support both critical public health research and outreach operations during a public health emergency. This success however, depended on community scholars’ leadership and integration with COVID-19 vaccination outreach–highlighting the synergy between community-engaged research and public health outreach operations. Importantly, the study also describes how this community-engaged program was developed and evolved in real-time during the pandemic. Finally, our study captures the Community Scholars’ own reflections, key insights, and contextualized strategies that, when combined with the operational description, could inform rapid deployment and refinement of community engagement methods for public health and healthcare in other crisis situations.

Our findings both contribute to and contrast with existing literature on Community-Based Participatory Research (CBPR). While CBPR emphasizes the necessity of building trust through academic-community partnerships, particularly in marginalized settings like meat processing plants, it also asserts the need for time to foster collaborative relationships that diminish pre-existing power imbalances between academia and communities.(16,18,18,19,21,46–49) However, public health emergencies such as mass COVID-19 outbreaks in large meat processing plants do not allow for the prerequisite time or optimal conditions. Early in the COVID-19 pandemic when little was known of SARS Cov-2‘s transmission dynamics, rapid research and concurrent public health and healthcare operations were urgently required to direct and coordinate health systems’ responses but were often difficult without pre-existing trust among migrant workers who faced significant structural barriers to healthcare and credible health information.(5,6,50) Despite these conditions, our study demonstrates the potential to rapidly launch and iteratively refine a CBPR program without pre-existing migrant worker engagement or relationships. Our study highlights the utility of CBPR as “action research”, wherein the purpose of research is not only to acquire knowledge, but to address unmet needs.(51–53) Our approach aligns with quality improvement literature that advocate starting and iteratively refining health system interventions to address identified issues than with traditional CBPR models that promote building trust first.(54) Our findings suggest that initiating urgent public health research and outreach, and trust building can occur simultaneously, indicating these concepts are not mutually exclusive and indeed most likely synergistic.

The pandemic exposed health inequities among racialized communities including migrant workers and underscored the urgent need to improve community engagement across high-income countries.(2–4,7,14,55,56) Innovative community-engaged healthcare and vaccine outreach programs that emerged during the pandemic successfully increased community trust, engagement, and vaccine uptake.(33,57,58) Nevertheless, these programs often depend on pre-existing partnerships and do not include research or data collection as a primary operational goal.(57) Unfortunately, lack of high quality health and health system data among socially marginalized migrant communities in high-income countries and their relative absence in health research, contribute to well described access and health outcome disparities between these communities and the general populations among host countries.(25,31,48) These persistent migrant health research gaps are not confined to high income countries alone. The WHO highlights how “[p]ersistent research gaps in these areas greatly impact the health of people who have migrated or been forcibly displaced and the health of communities worldwide.”(25) Our study provides evidence that extends the community-engagement paradigm by demonstrating the synergy of incorporating Community Scholars concurrently across participatory research and healthcare operations – greatly benefiting the objectives of both.

The Community Scholars’ reflections corroborate the synergies of their dual roles, revealing how their advocacy efforts were fueled by witnessing perceived injustices and how their research involvement ultimately led to empowerment to facilitate COVID-19 vaccine outreach and develop a deeper commitment to their communities.(33) These experiences motivated CSs to pursue further formal research training in the Patient and Community Engagement Research (PaCER) program,(45) underscoring the integrative potential of CBPR principles in practice-based research networks, and providing a real world example amidst a public health crisis.(18) These findings seem particularly important today, as trust in science since the pandemic has declined in many countries, and existing research points to community engaged partnerships with public health as key to rebuild lost trust(59,60)

Our study must be interpreted within the limitations of its methods and context. First, conducted in a single western Canadian province, the generalizability of our findings may be limited, though they likely resonate with similar sectors reliant on migrant labor. Second, while extensive, our inclusion of ethnocultural minority communities in Canada, is not exhaustive.

Despite this, the migrant communities represented in our study from Africa, Latin America, and Southeast Asia are similarly represented among migrant workers in other jurisdictions. Third, our community engagement methods were imperfect as our survey completion rate was 42.4% – potentially reduced by only utilizing an English version. Irrespective, this prompts further examination of engagement strategies. Finally, we cannot establish causality between CSs’ involvement in public health outreach operations and improved study recruitment among meat processing plant workers, the relationship is undoubtedly interdependent and warrants further exploration.

## 5. Conclusion

Our study contributes to the limited peer-reviewed literature on the lived experiences of migrant workers during the pandemic and illustrates the viability of successfully initiating and iteratively refining a community-based-participatory-research and outreach program amidst a public health emergency. Indeed, our findings indicate community engaged public health research and healthcare operations can begin, co-occur, and develop into meaningful community-academic-healthcare partnerships with minimal pre-existing infrastructure or institutional relationships to address critical public health research and operational needs simultaneously. Community scholars’ authentic engagement and pivotal role in these partnerships during the pandemic highlight their value in addressing public health challenges and may inform other health contexts to incorporate similar strategies to redress health inequities and produce much needed high-quality health research with migrant populations.

## Supporting information

GRAMMS Checklist

GRIPP2 Checklist

## Data Availability

All data produced in the present work are contained in the manuscript

## Conflict of Interest Statement

No competing interests declared.

## Funding Statement

This study was funded through the Canadian Institutes for Health Research (CIHR Application no. 469206)

## Supplementary Materials

**eTable 1:** Key COVID-19 Related Public Health Measures, Restrictions, & Relaunch Strategy in Alberta

## Acknowledgements

We would like to acknowledge the many newcomer communities who participated in this work and shared their experiences with us. We value your time and your trust, and hope that this work can inform the desired changes for newcomers.

## Author Contributions

GEF, KP, and EN led the conceptualization of this paper. MA, MY, AS, MY, and ERC actively supported data collection, data analysis, and manuscript preparation. EN, NH, OM, RG and AS provided support with writing, revising, and editing. MS, IN, AC, AB, SE, and DS provided, subject-matter expertise, and supported manuscript writing, revising, and editing. GEF and KP take responsibility for the final manuscript and provided scientific oversight throughout. All authors approved the final manuscript.

---

**eTable 1:**
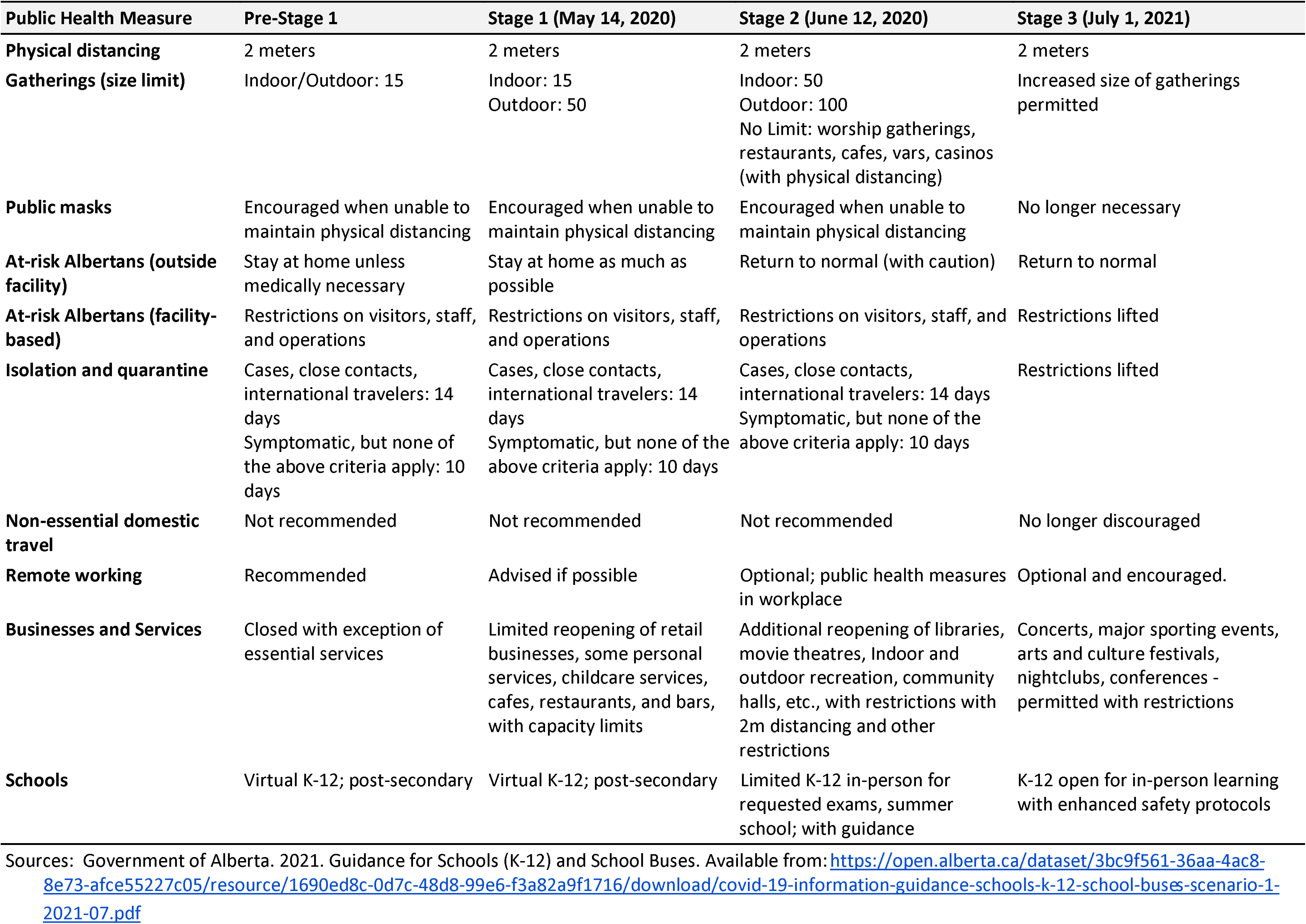
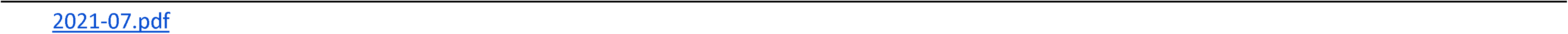
Key COVID-19 Related Public Health Measures, Restrictions, & Relaunch Strategy in Alberta.

| Public Health Measure | Pre-Stage 1 | Stage 1 (May 14, 2020) | Stage 2 (June 12, 2020) | Stage 3 (July 1, 2021) |
| --- | --- | --- | --- | --- |
| **Physical distancing** | 2 meters | 2 meters | 2 meters | 2 meters |
| **Gatherings (size limit)** | Indoor/Outdoor: 15 | Indoor: 15<br>Outdoor: 50 | Indoor: 50<br>Outdoor: 100<br>No Limit: worship gatherings, restaurants, cafes, vars, casinos (with physical distancing) | Increased size of gatherings permitted |
| **Public masks** | Encouraged when unable to maintain physical distancing | Encouraged when unable to maintain physical distancing | Encouraged when unable to maintain physical distancing | No longer necessary |
| **At-risk Albertans (outside facility)** | Stay at home unless medically necessary | Stay at home as much as possible | Return to normal (with caution) | Return to normal |
| **At-risk Albertans (facility-based)** | Restrictions on visitors, staff, and operations | Restrictions on visitors, staff, and operations | Restrictions on visitors, staff, and operations | Restrictions lifted |
| **Isolation and quarantine** | Cases, close contacts, international travelers: 14 days<br>Symptomatic, but none of the above criteria apply: 10 days | Cases, close contacts, international travelers: 14 days<br>Symptomatic, but none of the above criteria apply: 10 days | Cases, close contacts, international travelers: 14 days<br>Symptomatic, but none of the above criteria apply: 10 days | Restrictions lifted |
| **Non-essential domestic travel** | Not recommended | Not recommended | Not recommended | No longer discouraged |
| **Remote working** | Recommended | Advised if possible | Optional; public health measures in workplace | Optional and encouraged. |
| **Businesses and Services** | Closed with exception of essential services | Limited reopening of retail businesses, some personal services, childcare services, cafes, restaurants, and bars, with capacity limits | Additional reopening of libraries, movie theatres, Indoor and outdoor recreation, community halls, etc., with restrictions with 2m distancing and other restrictions | Concerts, major sporting events, arts and culture festivals, nightclubs, conferences - permitted with restrictions |
| **Schools** | Virtual K-12; post-secondary | Virtual K-12; post-secondary | Limited K-12 in-person for requested exams, summer school; with guidance | K-12 open for in-person learning with enhanced safety protocols |

Sources: Government of Alberta. 2021. Guidance for Schools (K-12) and School Buses. Available from: https://open.alberta.ca/dataset/3bc9f561-36aa-4ac8-8e73-afce55227c05/resource/1690ed8c-0d7c-48d8-99e6-f3a82a9f1716/download/covid-19-information-guidance-schools-k-12-school-buses-scenario-1-

## Notes

### Competing Interest Statement

The authors have declared no competing interest.

### Author Declarations

The study was approved by the University of Calgary research ethics board (REB20-1153)

