## Supplementary material for "A Community-Engaged Public Health Research and Outreach Program for Migrant and Racialized Workers in Meat Processing to Mitigate COVID-19 Inequities": GRAMMS Checklist

| Good Reporting of A Mixed Methods Study (GRAMMS) |  |
| --- | --- |
| Reporting Item | Where in Manuscript |
| (1) Describe the justification for using a mixed methods approach to the research question | Introduction: Page 3 |
| (2) Describe the design in terms of the purpose, priority and sequence of methods | Methods: Page 4-5 |
| (3) Describe each method in terms of sampling, data collection, and analysis. | Methods: Page 4-5 |
| (4) Describe where integration has occurred, how it has occurred, and who has participated in it. | Methods and Results: Page 5-9 |
| (5) Describe any limitation of one method associated with the present of the other method. | Discussion: Page 12 |
| (6) Describe any insights gained from mixing or integrating methods. | Discussion: Page 12 |

O'Cathain A, Murphy E, Nicholl J. The quality of mixed methods studies in health services research. J Health Serv Res Policy. 2008;13: 92-98.
