## Supplementary material for "A Community-Engaged Public Health Research and Outreach Program for Migrant and Racialized Workers in Meat Processing to Mitigate COVID-19 Inequities": GRIPP2 Checklist

| **Section and Topic** | **Item Reported** | **Page Number** |
| --- | --- | --- |
| Section 1: Abstract of paper | | |
| 1a: Aim | To engage migrant workers in meat processing industries in public health research and vaccine outreach using CBPR to mitigate COVID-19 related inequities. | 2 |
| 1b: Methods | CBPR methods were employed to recruit, train, and integrate community scholars who facilitated engagement and vaccine outreach among migrant workers. | 2 |
| 1c: Results | We recruited and trained six community scholars who then surveyed 191, and interviewed 43 workers in seven primary languages across eleven meat plants over three study phases between January 2021 and February 2022 | 2 |
| 1d: Conclusions | CBPR proved effective in enhancing engagement among migrant workers in both research and public health outreach. | 2 |
| 1e: Keywords | CBPR, COVID-19, meat processing workers, vaccine outreach, public health inequities. | 2 |
| Section 2: Background to paper | | |
| 2a: Definition | CBPR is defined as a collaborative approach to research that equitably involves community members, organizational representatives, and researchers in all aspects of the research process. | 3 |
| 2b: Theoretical underpinnings | The study is grounded in principles of participatory research, health equity, and inclusivity. | 4 |
| Section 3: Aims of paper | | |
| 3: Aim | To implement a CBPR approach in engaging racialized migrant workers in public health outreach and research during the COVID-19 pandemic. | 4 |
| Section 4: Methods of paper | | |
| 4a: Design | The study adopted a CBPR framework, engaging community scholars in designing, implementing, and evaluating the research and outreach activities. | 5 |
| 4b: People involved | Community scholars from various racialized ethnocultural groups, representing the migrant worker population. | 5 |
| 4c: Stages of involvement | Community scholars were involved in all stages, from research design to data collection, analysis, and dissemination. | 5 |
| 4d: Level or nature of involvement | Community scholars had a significant role in decision-making processes and were instrumental in tailoring outreach efforts to the community's needs. | 5 |
| Section 5: Capture or measurement of CBPR impact | | |
| 5a: Qualitative evidence of impact | Reflexivity activities and narrative analysis were used to qualitatively assess the impact of CBPR on the community scholars and the migrant workers. | 7-11 |
| 5b: Quantitative evidence of impact | Once a trusting relationship between community scholars and meat packing plant workers study registration notably improved. | Figure 1a |
| 5c: Robustness of measure | Community scholars and the research team used collective sense-making to review codes and finalize narrative analysis. | 6 |
| Section 6: Economic assessment | | |
| 6: Economic assessment | No specific economic assessment was completed during this study. |  |
| Section 7: Study results | | |
| 7a: Outcomes of CBPR | Enhanced trust and cooperation among migrant workers, leading to increased participation in vaccination programs and health research. | 6-11 |
| 7b: Impacts of CBPR | Positive impacts included improved public health outcomes and increased empowerment among community scholars and workers. | 6-11 |
| 7c: Context of CBPR | The context of urgent public health needs due to the pandemic highlighted the effectiveness and adaptability of CBPR in crisis situations. | 6-11 |
| 7d: Process of CBPR | The iterative process of engaging community scholars and refining strategies based on ongoing feedback was the core of this study. | 6-11 |
| 7ei: Theory development | The study demonstrated the viability and efficacy of implementing CBPR in emergency public health responses. | 6-11 |
| 7eii: Theory development | Not applicable. |  |
| 7f: Measurement | Figure 1a demonstrates the measured increase in participation over time. | Figure 1a |
| 7g: Economic assessment | Not applicable. |  |
| Section 8: Discussion and conclusions | | |
| 8a: Outcomes | CBPR significantly influenced the study by enhancing engagement and trust among migrant workers, leading to successful health interventions. | 11-12 |
| 8b: Impacts | The study underscores the value of CBPR in improving public health responses and highlights the need for its broader adoption in health research. | 11-12 |
| 8c: Definition | The use of CBPR as defined in this study aligns with existing literature and its application in this context validates its effectiveness. | 11-12 |
| 8d: Theoretical underpinnings | The study adds to the body of knowledge on the application of CBPR in emergency public health situations, supporting its theoretical underpinnings. | 11-12 |
| 8e: Context | The unique challenges of the COVID-19 pandemic provided a context that demonstrated the adaptability and effectiveness of CBPR. | 11-12 |
| 8f: Process | The three study phases represent three different processes: Scholar recruitment and training, early commmunity engagement and building trust, and community outreach vaccinations. | 11-12 |
| 8g: Measurement and capture of CBPR impact | Time series data and visualizations thereof were used to understand study recruitment over time. | Figure 1a |
| 8h: Economic assessment | Not applicable. |  |
| 8i: Reflections/critical perspective | The study was successful, however early methods to engage meat plant workers in research were ineffective as trust had not been fully established by that point. Future studies should see trust as a necessary criteria for CBPR. | 9 |
